# Validation of a Computerized Cognitive Test Battery for Detection of Dementia and Mild Cognitive Impairment

**DOI:** 10.1101/2020.11.10.20229369

**Authors:** Siao Ye, Kevin Sun, Duong Huynh, Huy Q. Phi, Brian Ko, Bin Huang, Reza Hosseini Ghomi

## Abstract

**Background:** Early detection of dementia is critical for intervention and care planning but remains difficult. Computerized cognitive testing provides an accessible and promising solution to address these current challenges. This study evaluated a computerized cognitive testing battery (BrainCheck) for its diagnostic accuracy and ability to distinguish the severity of cognitive impairment.

**Methods:** 99 participants diagnosed with Dementia, Mild Cognitive Impairment (MCI), or Normal Cognition (NC) completed the BrainCheck battery. Statistical analyses compared participant performances on BrainCheck based on their diagnostic group.

**Results:** BrainCheck battery performance showed significant differences between the NC, MCI, and Dementia groups, achieving ≥88% sensitivity/specificity for separating NC from Dementia, and ≥77% sensitivity/specificity in separating the MCI group from NC/Dementia groups. Three-group classification found true positive rates ≥80% for the NC and Dementia groups and ≥64% for the MCI group.

**Conclusions:** BrainCheck was able to distinguish between diagnoses of Dementia, MCI, and NC, providing a potentially reliable tool for early detection of cognitive impairment.

## Introduction

In proportion with the increasingly aging population, the incidence of dementia is on the rise; projected to affect nearly 14 million people in the United States and upwards of 152 million people globally in the coming decades [1–3]. Current rates of undetected dementia are reported as high as 61.7% [4], and available treatments are limited to promoting quality of life rather than reversal or cure of the disease process. The ability to properly identify and treat dementia at this scope requires an active approach focused on early identification. Early detection of dementia provides access to timely interventions and knowledge to promote patient health and quality of life before symptoms become severe. Early and accurate diagnosis also allows for proper preparation for patients, caregivers, and their families, resulting in improved caregiver well-being and delayed nursing home placements [5, 6]. Further, it helps to characterize early-stage dementia patients for clinical trials, exploring the latest therapeutics and validating biomarkers indicative of specific pathologies. Despite the benefits, early detection is a challenge with current clinical protocols, leaving many patients undiagnosed until symptoms become noticeable in later stages of the illness.

Considered an early symptomatic stage of dementia, mild cognitive impairment (MCI) signifies a level of cognitive impairment between normal cognition (NC) and dementia [7]. While not all MCI cases progress, the conversion rate of MCI to dementia has been observed at approximately 5%-10% [8]. This stresses the importance of identifying MCI in early detection and clinical intervention for dementia, included in recommendations from the National Institute on Aging and the Alzheimer’s Association [9]. Detection of MCI has been successful using brief cognitive screening assessments. The widely used Montreal Cognitive Assessment (MoCA) has demonstrated 83% sensitivity and 88% specificity in distinguishing MCI from NC, and 90% sensitivity and 63% specificity in distinguishing dementia from MCI [10, 11]. Similar performance has also been observed for the Mini-Mental State Examination (MMSE) and the Saint Louis University Mental Status (SLUMS) exam [12-14]. While these screening tests do well in their ability to detect MCI, they have many limitations. First, these tests are time- and labor-intensive (i.e. verbal administration by a physician or test administrator, hours for training, recording responses, scoring, and interpreting results). Second, these paper-based tests can not allow for tracking of timing, which are important indicators of an individual’s cognitive health. Also, there is a lack of detailed insights into different cognitive domains because their individual subtests are by design simple and suffer from ceiling effects [15-17].

Neuropsychological tests (NPTs) represent a more extensive and comprehensive class of cognitive evaluation. They allow for research into certain cognitive domains (e.g., attention, working memory, language, visuospatial skills, executive functioning, memory), that are used to support clinical diagnoses and further delineate specific neurocognitive disorders. NPTs can determine patterns of cognitive functioning that relate to normal aging, MCI, and dementia progression with a specificity of 67% − 99% [18]. A major strength of NPTs is the ability to characterize cognitive impairment, providing clues to underlying pathology, and thereby improving diagnostic accuracy to guide appropriate treatment. However, NPTs come with downsides including financial cost, long appointment times, and high levels of training and expertise required to conduct and interpret tests. Prior studies have also shown that some NPTs demonstrate high accuracy in differentiating dementia patients from healthy participants, but do not have adequate psychometrics to distinguish MCI from dementia [19–22].

Computerized cognitive assessment tools have been developed to address the issues of accessibility and efficiency [23, 24]. They are more comprehensive than screening tests but less expensive and quicker than clinical NPTs, aimed to maximize accessibility to both patients and providers. This would also yield multiple benefits including maintaining testing standardization, alleviating the time pressures of modern clinical practice, and providing a comprehensive assessment of cognitive function to strengthen a clinical diagnosis. Importantly, in the new era of practicing amid the COVID-19 pandemic [25], increasing the accessibility of remote cognitive testing for vulnerable/high-risk patients is essential.

This study evaluated BrainCheck, a computerized cognitive test battery available on mobile devices such as smartphones or tablets, that is portable and able to be administered remotely. In addition to offering automated scoring and instant interpretation, BrainCheck requires short administration and testing time, comparable to traditional screening instruments, but provides detailed insight into multiple aspects of cognitive functioning that only comprehensive NPT can. BrainCheck has previously been validated for its diagnostic accuracy in detecting concussion [26] and dementia-related cognitive decline [27]. Furthering its validation for dementia-related cognitive decline, we sought to assess BrainCheck’s utility as a diagnostic aid to accurately assess the severity of cognitive impairment. We measured BrainCheck’s ability to distinguish individuals with different levels of cognitive impairment (NC, MCI, and Dementia) based on their comprehensive clinical diagnoses. Our goal was to further demonstrate the utility of BrainCheck for cognitive assessment, specifically as a diagnostic aid in cases where NPT may be unavailable, or when a comprehensive evaluation is not indicated.

## Methods

### Recruitment

This study was approved by the University of Washington (UW) Institutional Review Board for human subject participation. Participants were recruited from a research registry maintained by the Alzheimer’s Disease Research Center associated with the UW Medicine Memory and Brain Wellness Center Clinic [28]. This registry is a continually updated database of individuals who have expressed interest and signed an IRB-approved consent form to be contacted about participation in Alzheimer’s Disease and Related Dementias research studies, many who have been recently evaluated at the clinic, and hence have a clinical diagnosis or evaluation. Those with listed addresses within a 70-mile radius of Seattle, Washington were contacted by phone or email provided within the registry. If the person was unable to physically use an iPad, too cognitively impaired to understand or follow instructions, or if the primary contact (e.g., spouse) indicated that the person was unable to participate, they were not recruited for the study. When study procedures were modified from in-person to remote administration due to the COVID-19 pandemic (approximately March 2020), participants outside the initial geographical range were contacted to explore remote testing capabilities. We required that these participants had access to either an iPad with iOS 10 or later, or a touchscreen computer and Wi-Fi connectivity to participate in the study.

Using the provided primary cognitive diagnosis within the registry, participants were divided into one of three groups: 1) Normal Cognition (NC), indicated by subjective cognitive complaint or no diagnosis of cognitive impairment, some of which were self-reported, 2) Mild Cognitive Impairment (MCI), representing both amnestic and non-amnestic subtypes, or 3) Dementia, which included dementia due to Alzheimer’s Disease (AD), Frontotemporal dementia, Vascular dementia, Lewy body dementia, Mixed dementia, or Atypical AD.

Five participants were not recruited from the registry but via snowball sampling from other participants. The recruitment of these participants was simply due to convenience, typically a family member or friend that was also available at the time of testing. Four of these participants were placed into the NC group after self-report denying symptoms or a history of cognitive impairment; these four were not patients of the memory clinic. One of the participants recruited from snowball sampling was a patient from the memory clinic, just not a part of the registry, and placed in the AD group based on their most recent diagnosis retrieved from their medical records. All five were on-site administration.

## Study design and procedures

### On-site administration

Data collection for on-site administration was collected from October 2019 to February 2020. A session was held either in the participant’s home or in a public setting that was well-lit, quiet, and distraction-free. Consent forms were reviewed and signed by the participant, or legally authorized representative, and examiner, with both parties obtaining a copy. The study was designed for participants to complete one session with a moderator using a provided iPad (Model MR7G3LL/A) connected to Wi-Fi to complete the BrainCheck battery. Prior to testing, participants were briefed on BrainCheck, and moderator guidance was limited to questions and assistance requested by the participant during the practice portions. Participants received a gift card for participation at the conclusion of the study session.

### Remote administration

Due to the COVID-19 pandemic and interest in preliminary data on remote cognitive testing, study procedures were modified to accommodate stay-at-home orders in Washington state. Data collection resumed from April 2020 to May 2020, with modified procedures using remote administration. These participants provided written and verbal consent and were administered the BrainCheck battery remotely over a video call with the moderator. Participants used their personal iPads or touchscreen computer browser to complete the BrainCheck battery. The same method for on-site administration (described above) was used for remote administration.

### Measurements

A short description of each of the 5 assessments comprising the BrainCheck battery (V4.0.0) are listed in Table 2. More detailed descriptions may be found in a previous validation study [26]. After completion of the BrainCheck battery, the score of each assessment is calculated using assessment-specific measurements by the BrainCheck software (Table 2). The BrainCheck Overall Score is a single, cumulative score for the BrainCheck battery that represents general cognitive functioning. This score is calculated by taking the average of all completed assessment scores. If an assessment is timed-out, a penalty is applied by setting this assessment score to be zero. The normalized assessment scores and BrainCheck Overall Scores are corrected for participant age and device used (iPad versus computer) using the mean and standard deviation of the corresponding score from a normative database previously collected by BrainCheck [26,27]. The score generated follows a standard normal distribution, where a lower score indicates lower assessment performance and cognitive functioning.

**Table 1.**
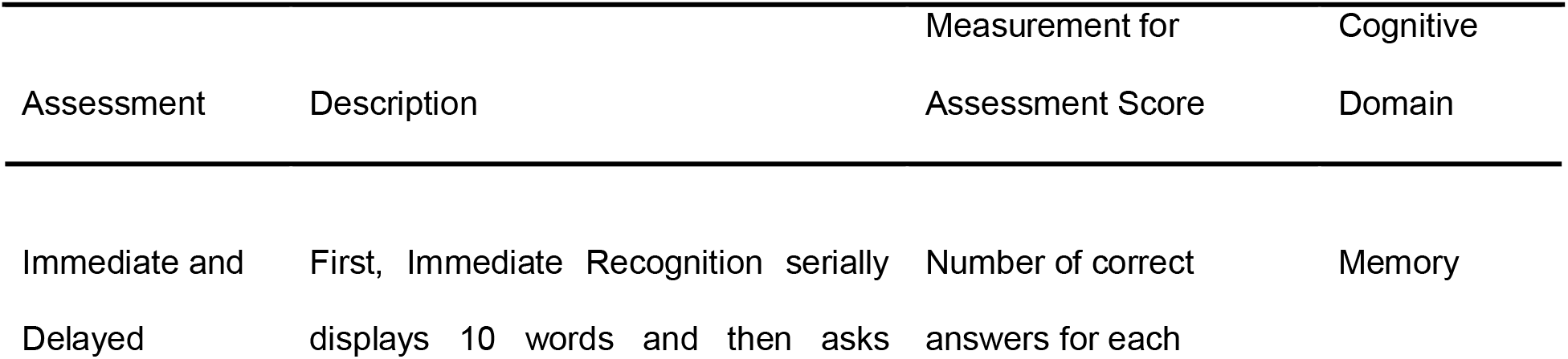
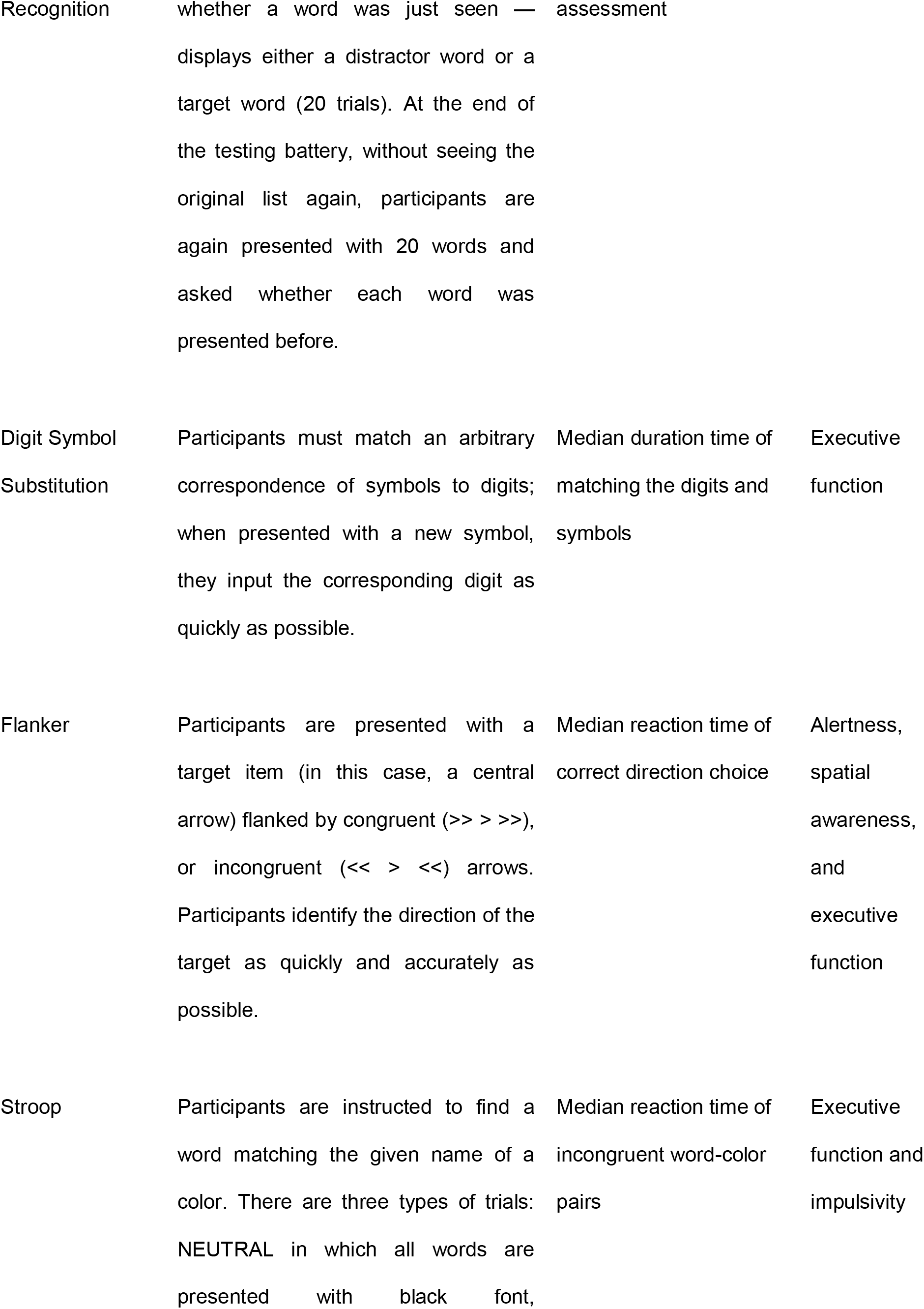
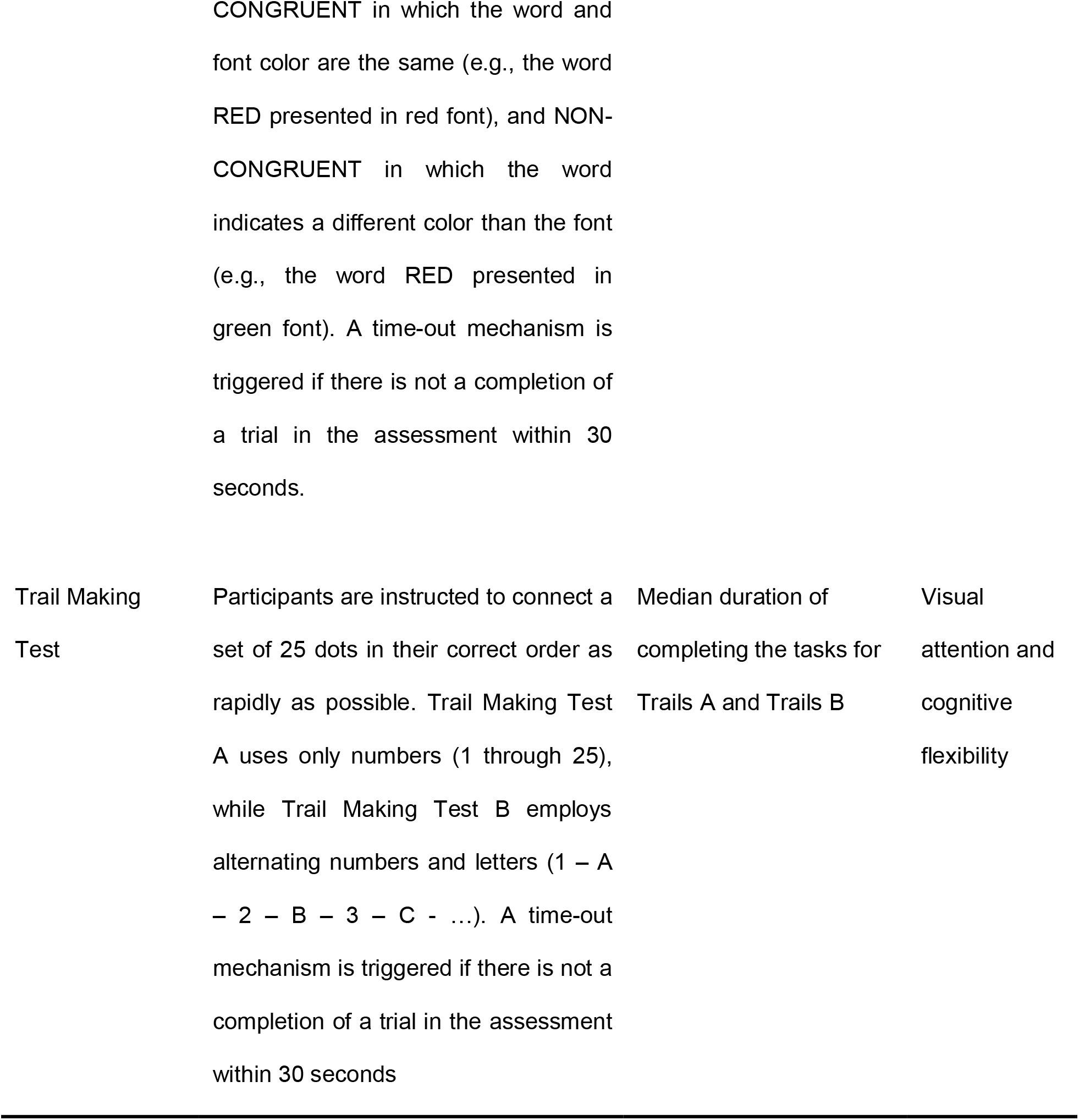
Cognitive assessments in BrainCheck.

**Table 2.** Participant demographics.

| Demographics | NC | MCI | Dementia | P-value |
| --- | --- | --- | --- | --- |
| *Sample (n = 99)* | 35 | 22 | 42 | - |
| *Age mean(SD)* | 67.8 (9.6) | 73.5 (5.9) | 71.5 (9.0) | **.04<sup>a</sup>** |
| *Sex* |  |  |  |  |
| Female | 25 | 8 | 16 | **<.005<sup>b</sup>** |
| Male | 10 | 14 | 26 |  |
| *Education level* |  |  |  |  |
| Some college or less | 2 | 2 | 8 | **.70<sup>b</sup>** |
| BA / BS college graduate | 10 | 6 | 11 |  |
| Post Bachelor | 14 | 9 | 16 |  |
| N/A | 9 | 5 | 7 |  |
| *Administration Type* |  |  |  |  |
| On-site | 29 | 16 | 29 | **.37<sup>b</sup>** |
| Remote | 6 | 6 | 13 |  |
| *Recruitment Type* |  |  |  |  |
| Registry | 31 | 22 | 41 | **.09<sup>b</sup>** |
| Snowball | 4 | 0 | 1 |  |

Significant p-values are shown in bold
<sup>a</sup> Calculated by the ANOVA test
<sup>b</sup> Calculated by the chi-square test

### Statistical analysis

Statistical analyses were performed using Python version 3.8.5 and R version 3.6.2 programming languages. All tests were two-sided, and significance was accepted at the 5% level (alpha = .05). Comparison of means of groups was made by an analysis of variance (ANOVA) test for normally distributed data. The chi-square test was used to analyze differences in categorical variables.

To evaluate BrainCheck performance among participants in different diagnostic groups while adjusting for age, sex, and administration type, linear regression was used in which the outcome variables were duration to complete BrainCheck battery, individual BrainCheck assessment scores, and BrainCheck Overall Scores. *P*-values were corrected using Tukey’s method for multiple comparisons. To assess the accuracy of the BrainCheck Overall Score in the binary classification of participants in the different diagnostic groups (NC vs. Dementia, NC vs. MCI, MCI vs. Dementia), receiver operating characteristic (ROC) curves with area under the curve (AUC) calculations were generated to determine diagnostic sensitivity and specificity. In assessing BrainCheck for three group classification, we used volume under the three-class ROC surface (VUS) method from Luo et al. [29], to define optimal cutoffs for the BrainCheck Overall Score and find the maximum diagnostic accuracy.

## Results

### Participant characteristics and demographics

A total of 241 individuals were contacted to participate and 99 participants completed the study. Demographic details of the participants are provided in Table 2. The three groups did not differ to a significant degree in terms of education, administration type, or recruitment type, but there were differences in age and sex.

### Completion of Assessments

We found most participants in the NC group were able to complete the assessments whereas the Dementia group had a higher ‘timeout’ rate, with MCI falling in between the two (Fig. 1). The timeout function occurs when a participant can not complete a trial of the assessment in 30 seconds; it is embedded in the Stroop and Trails A/B assessments. Timeouts were mainly due to response delays; where participants were attempting the test but could not answer quickly enough. Overall, the Dementia group took significantly more time to complete the BrainCheck battery (*median*=30.5 mins, *IQR*=23.4-37.1 mins) compared to the MCI (*median*=21.5 mins, *IQR*=19.3-24.2 mins) and NC group (*median*=17.8 mins, *IQR*=15.4-19.6 mins).

**Fig. 1:**
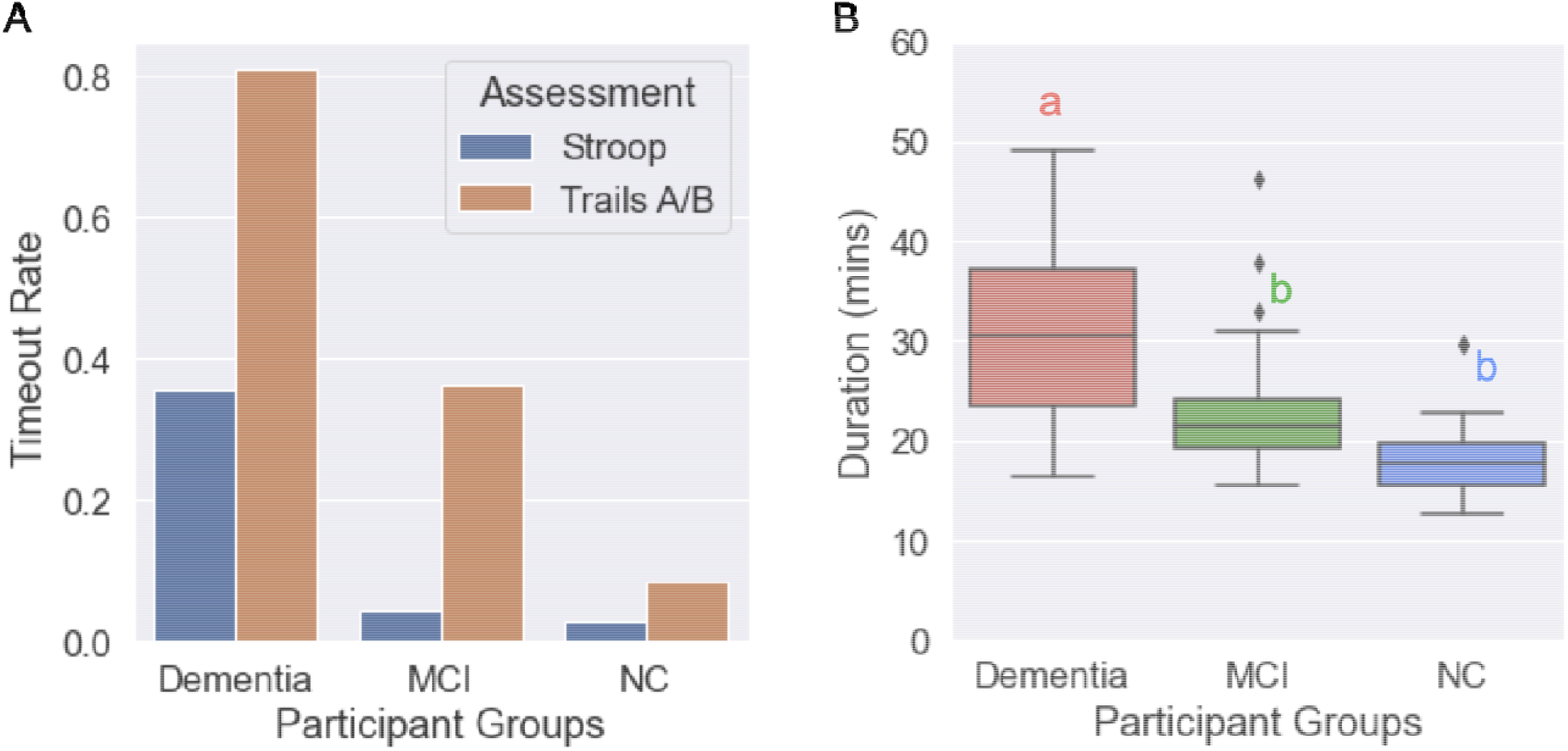
Completion of assessments and durations to complete BrainCheck battery. (A) Timeout rates of the Stroop and Trails A/B assessments for each diagnostic group. The BrainCheck Stroop and Trails A/B assessments timeout if participants cannot complete a trial of the assessment in 30 seconds. (B) Duration (mins) to complete the BrainCheck Battery for each diagnostic group. Letters (*a, b*) indicate significant differences between groups (*P*<.05) in the linear regression model adjusted for age, gender, and administration type; any two groups sharing a letter are not significantly different.

### BrainCheck performance

BrainCheck assessments were compared across the three groups using a linear regression model adjusted for age and gender (Fig. 2 and Table 3). Individual scores, such as the BrainCheck Overall Score, were normalized for age and device. Overall, participants with greater cognitive impairment showed lower BrainCheck assessment scores. All individual assessments except Trails B showed significant differences in performance between the NC and Dementia groups, while two of the seven assessments (Immediate Recognition and Digit Symbol Substitution) showed significant differences in performance between all three groups (Fig 2 and Table 3). Digit Symbol Substitution, Flanker, and Trails A/B assessments showed long tails in the scores of the Dementia group because some participants in the Dementia group only completed parts of the assessments or exhibited low accuracy (Fig SI-1).

**Table 3.** Linear regression model analyses using each BrainCheck assessment score and BrainCheck Overall Score as the outcome variable in separate adjusted models^1^.

|  | NC | MCI | Dementia | Dementia<br>vs. NC | Dementia<br>vs. MCI | MCI vs. NC |
| --- | --- | --- | --- | --- | --- | --- |
| Assessments | Estimated Marginal Means (SE) |  |  | Contrast Estimate (*P*-value) |  |  |
| Immediate | .17 | -1.93 | -3.36 | -3.54 | -1.43 | -2.10 |
| Recognition* | (.44) | (.50) | (.36) | **(<.001)** | **(.04)** | **(.005)** |
| Delayed | .06 | -2.16 | -2.92 | -2.98 | -.76 | -2.23 |
| Recognition | (.34) | (.39) | (.28) | **(<.001)** | (.23) | **(<.001)** |
| Digit Symbol | .80 | -.21 | -1.23 | -2.02 | -1.01 | -1.01 |
| Substitution* | (.27) | (.29) | (.29) | **(<.001)** | **(.04)** | **(.03)** |
| Flanker | .76 | -.74 | -2.64 | -3.4 | -1.89 | -1.5 |
|  | (.45) | (.51) | (.41) | **(<.001)** | **(.009)** | (.06) |
| Stroop | -.43 | -.63 | -.91 | -.49 | -.28 | -.21 |
|  | (.12) | (.13) | (.12) | **(.01)** | (.23) | (.46) |
| Trails A | -.01 | -.75 | -1.69 | -1.67 | -.94 | -.74 |
|  | (.33) | (.36) | (.30) | **(<.001)** | (.11) | (.29) |
| Trails B | .51 | .21 | -.16 | -.67 | -.37 | -.30 |
|  | (.21) | (.23) | (.24) | (.08) | (.47) | (.57) |
| Normalized | .71 | -2.15 | -5.63 | -6.34 | -3.48 | -2.86 |
| BrainCheck | (.55) | (.62) | (.45) | **(<.001)** | **(<.001)** | **(.002)** |
| Overall Score* |  |  |  |  |  |  |

Significant p-values are shown in bold. Assessments marked with \* indicate significant differences across all three diagnostic groups.
<sup>1</sup> Adjusted for age, sex, and administration type

**Fig. 2:**
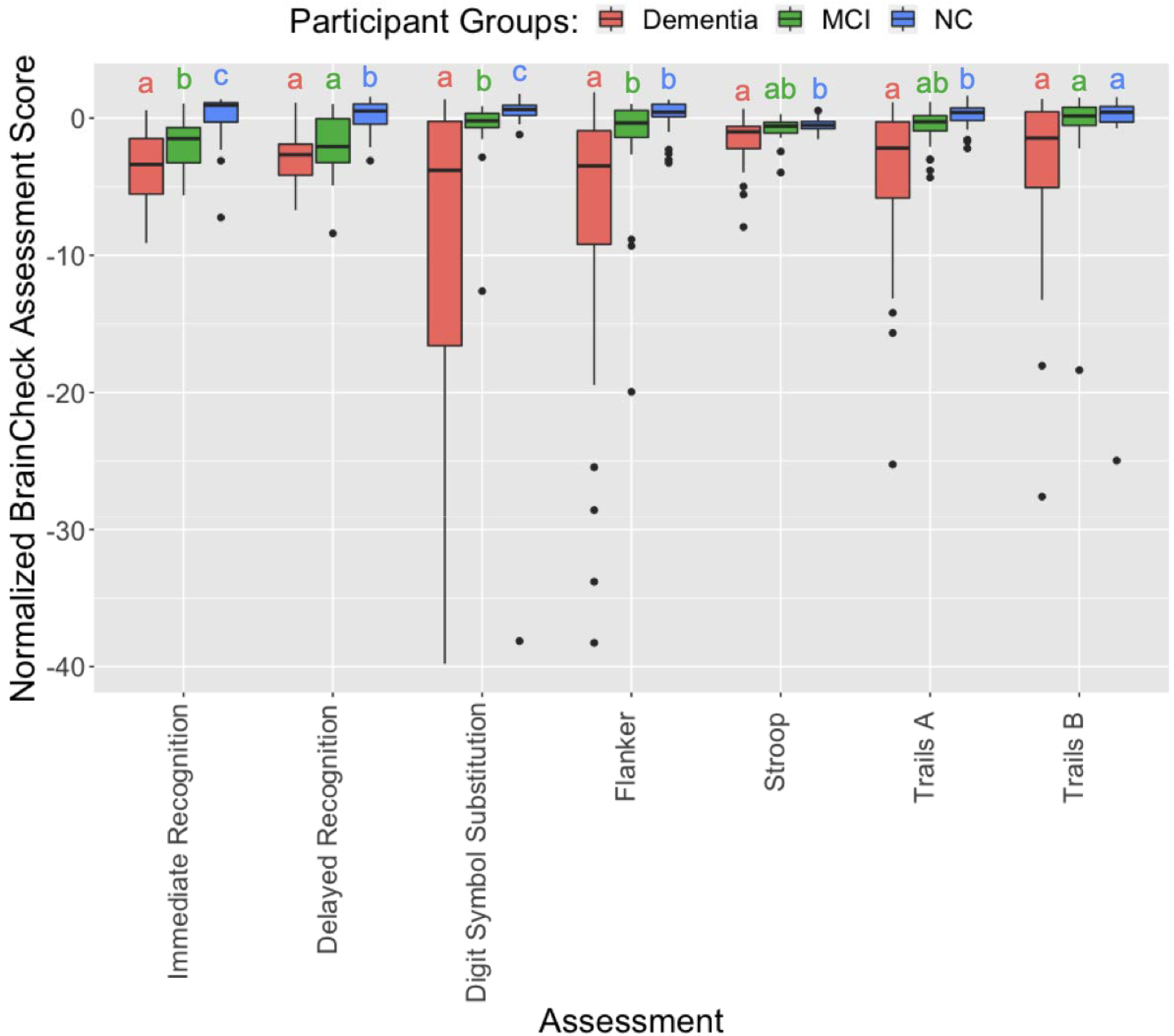
Pairwise comparison of participant groups based on normalized scores of BrainCheck assessments. For each assessment, any two groups sharing a letter are not significantly different. Otherwise, they are significantly different (*P*<.05) in linear regression models adjusted for age, gender, and administration type. The outliers identified by the IQR method in each assessment are removed before the comparison.

The BrainCheck Overall Score is a composite of all individual assessments within the BrainCheck battery, representing overall performance (See details under the measurements section). Using an existing normative population database, partly compiled from controls in previous studies [24,25], the BrainCheck Overall Score was adjusted for age and the device used to generate the normalized BrainCheck Overall Scores. The normalized BrainCheck Overall Scores differed significantly among these three groups (*P*<.05). The NC group scored significantly higher than the MCI and Dementia groups. The Dementia group demonstrated significantly lower scores than the other two groups (Fig. 3).

**Fig 3:**
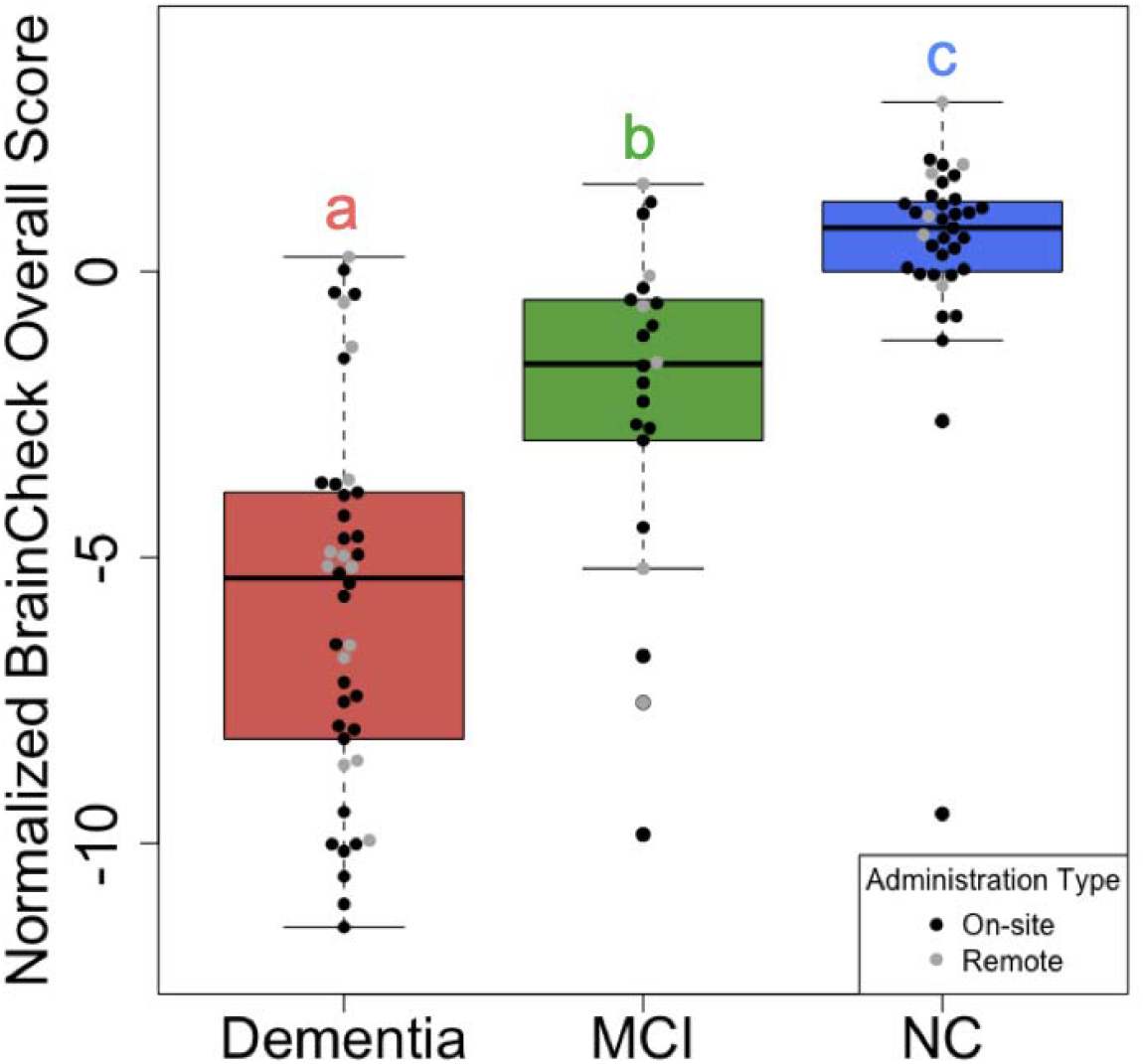
Comparison of normalized BrainCheck Overall Scores among groups. The normalized BrainCheck Overall Score follows a standard normal distribution. Letters (*a, b, c*) indicate significant differences (*P*<.05) on the linear regression model adjusted for age, gender, and administration type.

### BrainCheck diagnostic accuracy

Using ROC analysis, Braincheck Overall Scores achieved a sensitivity of 94% and a specificity of 88% for classifying NC and Dementia participants (AUC=.95); a sensitivity of 83% and a specificity of 86% for classifying between NC and MCI participants (AUC=.84); and a sensitivity of 77% and a specificity of 83% for classifying between MCI and Dementia participants (AUC=.79) (Fig 4). Using methods described by Luo and colleagues for three group classification [29], the optimal lower and upper cutoffs of the normalized BrainCheck Overall Score in maximizing diagnostic accuracy were −3.64 and −.06, respectively. This achieved true positive rates of 83% for the NC group, 64% for the MCI group, and 83% for the Dementia group (Fig. 5).

**Fig 4.**
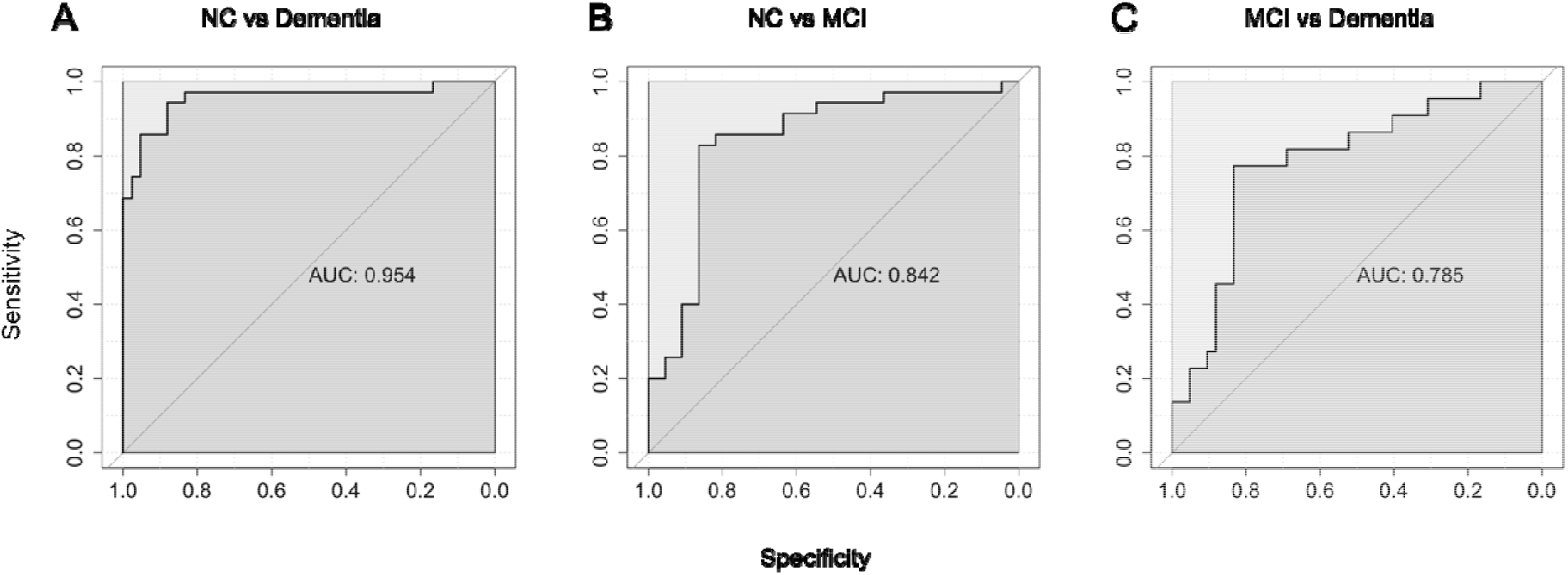
ROC curves for the BrainCheck Overall Score in classifying participants of different groups. ROC curves with AUC’s for the BrainCheck Overall Score in the binary classification of (A) NC vs. Dementia, (B) NC vs. MCI, (C) MCI vs. Dementia.

**Fig. 5:**
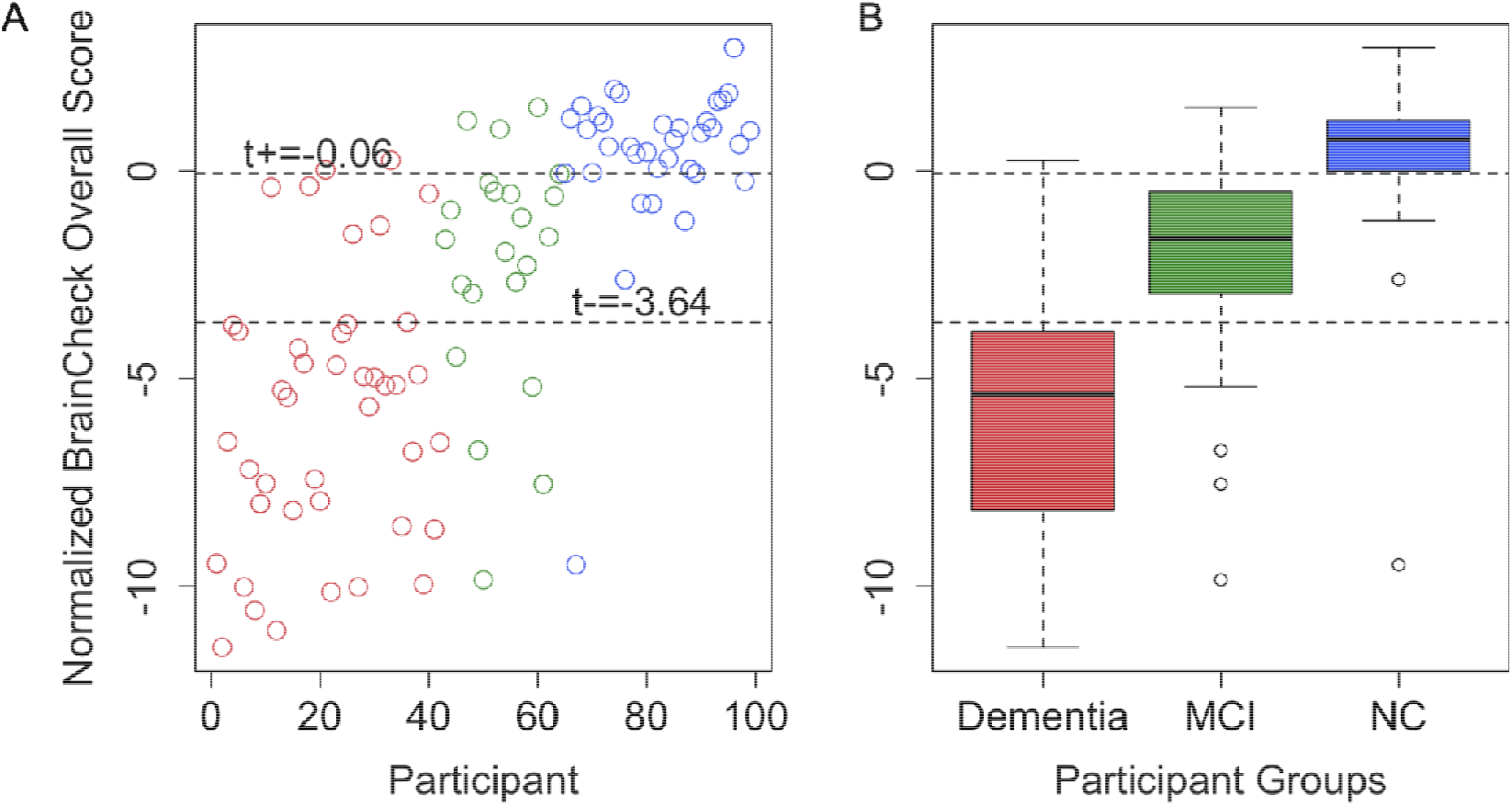
Optimal BrainCheck cutoff scores for distinguishing NC, MCI, and Dementia groups. (A) Individual participant normalized BrainCheck Overall Scores, where the x-axis is the index of the participant, sorted by primary diagnosis (Dementia - red, MCI - green, and NC - blue). (B) Boxplots of normalized BrainCheck Overall Scores for each diagnostic group. The normalized BrainCheck Overall Score follows a standard normal distribution. The dashed lines label the optimal cutoff scores for distinguishing the diagnostic groups.

## Discussion

Consistent with prior findings in concussion [26] and dementia/cognitive decline [27] samples, this study demonstrated that BrainCheck is consistent in its capability to detect cognitive impairment, and can reliably detect and differentiate between the severity of cognitive impairment groups (NC, MCI, and Dementia). As expected, participants with more severe cognitive impairment performed worse across the individual assessments and on BrainCheck Overall Scores. The BrainCheck Overall Scores separated participants of different diagnostic groups successfully with high sensitivity and specificity.

BrainCheck Overall Scores were more robust in distinguishing these groups where participants in the Dementia group had significantly lower scores than ones in the NC group. The BrainCheck battery was able to distinguish between NC and Dementia participants, with 94% sensitivity and 88% specificity. These findings show that the BrainCheck Overall Score demonstrates better accuracy for differentiating NC from dementia, compared to the MMSE, SLUMS, and MoCA screening measures [14, 29, 31]. People with MCI usually experience fewer cognitive deficits and preserved functioning in activities of daily living compared to those with dementia [32], and our findings of sensitivity and specificity with separating MCI from other groups were slightly lower than the NC versus the Dementia differentiations (Fig. 5). Nonetheless, the BrainCheck Overall Score showed sensitivities and specificities greater than 80% in distinguishing MCI from NC and Dementia groups, which is comparable to the MoCA, SLUMS, and MMSE [11-14, 31]. Furthermore, a review of validated computerized cognitive tests indicated AUC’s ranging from .803 to .970 for detecting MCI, and AUCs of .98 and .99 in detecting dementia due to AD [30], which were comparable with the results found in this study.

Although not all individual assessments in the BrainCheck battery differentiated between NC, MCI, and Dementia, we observed a general trend for each assessment showing that Dementia participants had the lowest scores whereas the NC participants had the highest scores. Individual assessments that did show significant differences in the scores between NC and MCI groups and between Dementia and MCI groups included the Immediate Recognition and Digit Symbol Substitution. Notably, the BrainCheck Digit Symbol test showed significant differences in performance between all three diagnostic groups, whereas a previous study found that the BrainCheck Digit Symbol test did not show significant differences between cognitively healthy and cognitively impaired groups (n=18, *P*=.29) [27]; likely due to this study having a larger sample size. Individual assessments with no significant differences between the MCI group and the NC/Dementia groups were the BrainCheck Stroop and Trails A/B tests (Fig 2 and Table 3). All of these tests include timeout mechanisms if participants are unable to complete the test, and time-out rates were higher in the more cognitively impaired groups (Fig 2). Therefore, when calculating the BrainCheck Overall Score, we have introduced a penalty mechanism for timed-out assessments.

In comparison to comprehensive NPT, which can typically last a few hours and sometimes require multiple visits [32], BrainCheck demonstrated shorter test duration, with median completion times of 17.8 min (*IQR*=15.4-19.6 min) for NC participants and 30.5 min (*IQR*=23.4-37.1 min) for dementia participants (Fig. 1). Shorter test durations observed in individuals with no/less cognitive impairment suggest that computerized cognitive tests could be useful for rapid early detection in this population, prompting further evaluation, whereas those with dementia have likely already undergone a comprehensive evaluation. The wide variance in completion time for the Dementia group may have uncovered the difficulty participants with more severe cognitive impairment may have faced in completing the BrainCheck battery, compared to the lower variance observed in the NC group.

A limitation of this study was that participants were not diagnosed by a physician at the time point of BrainCheck testing. Thus, participants were placed into the diagnostic groups based on their most recent clinical diagnosis available in their electronic health record, or for the few NC participants without medical evaluations, based on their report of no cognitive symptoms or diagnosis or cognitive impairment. The period from the most recent clinical diagnosis to the date of BrainCheck testing varied among the diagnostic groups; the Dementia group had the fewest days from their latest clinical evaluation (*median*=82.5 days; *IQR*=44.5-141.25), followed by the MCI group (*median*=244 days, *IQR*=105-346.5) and the NC group (*median*=645 days; *IQR*=225.5-1112.5). These large time intervals in a degenerative population leave room for cognition to worsen over time, potentially blurring the lines in the severity of cognitive impairment, where participants may have progressed to MCI from NC, and to Dementia from MCI during that period. This would make the distinguishing of NC from cognitive impairment more difficult, yet diagnostic accuracy among the groups remains high. Furthermore, the median number of days since the last clinical evaluation for NC participants was as high as 645 days. This could suggest that the NC participants did not feel an inclination to seek out further cognitive evaluation during the extended time period, and may not have experienced noticeable cognitive decline. Future validity studies should ensure that a physician evaluation and diagnosis occur closer to the time of BrainCheck testing to address these limitations.

Another limitation was that, although not all individual assessment scores could differentiate the three groups, the pattern of differences across these scores may contain useful diagnostic information. The use of the BrainCheck Overall Score as an average of all individual assessment scores appears to work effectively, but does not take into account the other relationships seen across individual scores. Furthermore, some individual scores may be more informative for detecting cases while others may be informative for gauging severeness. Future studies recruiting a larger sample size in each group will allow for an investigation into whether machine learning methods can extrapolate these relationships and improve the diagnostic accuracy of BrainCheck.

When administration type was considered in linear regression model analyses, scores only showed significant differences among the three diagnostic groups instead of administration types. While remote administration was not designed into the original study, stay-at-home orders due to COVID-19 required modifications, and efforts were made to provide preliminary data for remote use. With preliminary outcomes indicating feasibility for remote administration, a more robust study and increased sample size will be needed to fully validate BrainCheck’s cognitive assessment via its remote feature.

## Conclusions

The use of computerized cognitive tests provides the opportunity to increase test accessibility for an aging population with an increased risk of cognitive impairment. Technological advancements have allowed these tools to obtain greater sensitivity and specificity compared to their paper and pen counterparts [30], and remote testing could bring a more time and cost-effective solution. The findings in this study demonstrate that BrainCheck could distinguish between three levels of cognitive impairment, NC, MCI, and Dementia. BrainCheck is automated and quick to administer, both in-person and remote, which could help increase accessibility to testing and early detection of cognitive decline in an ever-aging population. This study paves the way for a comprehensive longitudinal study, exploring BrainCheck in early detection of dementia and monitoring of cognitive symptoms over time, including further comparison to ‘gold standard’ neuropsychological assessments.

## Data Availability

Data may be made available by contacting the corresponding author with a data use agreement.

https://depts.washington.edu/mbwc/adrc/page/research-resources

## Abbreviations

MCI: Mild Cognitive Impairment
NC: Normal Cognition
MoCA: Montreal Cognitive Assessment
SLUMS: Saint Louis University Mental Status
MMSE: Mini-Mental State Examination
NPT: Neuropsychological Testing
AD: Alzheimer’s Disease
ANOVA: Analysis of Variance
ROC: Receiver Operating Characteristic
AUC: Area Under the Curve
UW: University of Washington

## Acknowledgments

Funding was provided by BrainCheck, Inc. Registry was provided by an NIH-funded research resource center associated with the University of Washington Memory and Brain Wellness Center and the University of Washington Alzheimer’s Disease Research Center. We would like to thank Carolyn M. Parsey, neuropsychologist at the University of Washington Memory and Brain Wellness Center at Harborview and a UW Assistant Professor of Neurology, for advising throughout the entirety of this study.

## Author contributions

Conceptualization: Bin Huang, Reza Hosseini Ghomi

Formal analysis: Siao Ye, Kevin Sun, Bin Huang

Funding acquisition: Reza Hosseini Ghomi

Investigation: Kevin Sun, Huy Phi, Brian Ko

Methodology: Siao Ye, Bin Huang, Reza Hosseini Ghomi

Resources: Kevin Sun, Reza Hosseini Ghomi

Supervision: Bin Huang, Reza Hosseini Ghomi

Writing – original draft: Siao Ye, Kevin Sun, Huy Phi, Brian Ko, Bin Huang

Writing – review & editing: Siao Ye, Kevin Sun, Duong Huynh, Huy Phi, Brian Ko, Bin Huang, Reza Hosseini Ghomi

## Conflicts of Interest

All authors report personal fees from BrainCheck, outside the submitted work; DH, BH, RHG report receiving stock options from BrainCheck. The study was funded by BrainCheck.

## Notes

### Competing Interest Statement

The following authors declare the following competing interests: BH, KS, RHG, SY, HP, BK report personal fees from BrainCheck, outside the submitted work; BH, RHG reports receiving stock options from BrainCheck.

### Author Declarations

Approval was granted with the University of Washington Human Subjects Division under study ID: STUDY00000790 on August 1st, 2019.

